# Beyond Haploinsufficiency: Protein Domain and Variant Class Shape Language Outcomes in SYNGAP1-DEE

**DOI:** 10.64898/2026.11.04.26367250

**Authors:** L. Kiwull, V. Schmeder, M. Zenker, M. Mengual Hinojosa, G. Kluger, K. Eschermann

## Abstract

**Background:** SYNGAP1-related developmental and epileptic encephalopathy (SYNGAP1-DEE) is generally considered a haploinsufficiency disorder, yet language development varies considerably between affected individuals. With targeted therapies, including antisense oligonucleotides (ASOs), approaching clinical evaluation, understanding genotype-specific differences is increasingly important. We investigated whether expressive language and functional communication are associated with SynGAP protein domains and variant class.

**Methods:** We analyzed cross-sectional caregiver-reported data linked to genetic reports from the international PATRE SYNGAP1 registry. Of 440 participants, 334 had variants that could be assigned to a protein domain and classified as missense (n=79) or truncating/splicing (n=255). Variants were mapped to seven predefined SynGAP protein regions. The primary outcome was a five-level current expressive language score. Secondary outcomes included acquisition of two-word phrases and CFCS-derived functional communication. Each domain was compared with all other regions within the same variant class using nonparametric tests and age-adjusted ordinal or logistic regression. Benjamini-Hochberg false discovery rate correction was applied.

**Results:** Missense variants in the C2 domain (aa248-392; n=28) were associated with markedly poorer expressive language (median score 1 vs. 2; age-adjusted OR 0.16, 95% CI 0.06-0.46; q=0.0025). Only 7.1% had acquired two-word phrases compared with 52.0% of individuals with missense variants outside C2 (OR 0.07; q=0.0023). Functional communication was also significantly impaired (OR 0.23, 95% CI 0.09-0.56; q=0.0052). Conversely, truncating/splicing variants in the PH domain (aa103-247; n=50) were associated with better expressive language (median 3 vs. 2; OR 3.52, 95% CI 1.99-6.24; q<0.001) and more frequent acquisition of two-word phrases (62.0% vs. 31.5%). No corresponding language association was observed for C2 truncating/splicing variants or for variants in the RasGAP domain.

**Conclusions:** Language outcomes in SYNGAP1-DEE are associated with both variant class and protein location. The marked impairment associated with C2 missense variants contrasts with the relatively preserved language abilities observed in PH truncating/splicing variants, suggesting that SYNGAP1-related disease cannot be fully explained by a uniform haploinsufficiency model alone. These findings support considering protein domain and variant class in future ASO trials, particularly for patient stratification, endpoint selection, and interpretation of treatment outcomes.

**Abstract Summary:** Language outcomes in SYNGAP1-DEE differ according to protein domain and variant class. C2 missense variants are associated with markedly poorer expressive language and functional communication, whereas PH truncating/splicing variants are linked to better language abilities. These findings may inform patient stratification and outcome assessment in upcoming antisense oligonucleotide trials.

## 1. Introduction

*SYNGAP1*-related developmental and epileptic encephalopathy (*SYNGAP1*-DEE) is a genetic neurodevelopmental disorder characterized by intellectual disability, developmental delay, epilepsy, and a broad range of behavioral difficulties [4,9]. One of its most striking features is the considerable variability in language development. While some affected individuals acquire functional speech and develop sentence-level communication, others remain limited to a few words or do not develop meaningful spoken language at all. Communication difficulties can substantially affect everyday functioning, independence, and social participation. Understanding the factors underlying this variability is therefore important for clinical counseling, assessment of developmental outcomes, and the design of future therapeutic studies [1,5,11].

The *SYNGAP1* gene encodes SynGAP, a synaptic GTPase-activating protein that plays a central role in glutamatergic synaptic signaling and plasticity. SynGAP is highly enriched in the postsynaptic density, where it regulates Ras- and Rap-dependent signaling pathways involved in synaptic maturation and AMPA receptor trafficking [2,3]. Pathogenic *SYNGAP1* variants are generally considered to cause disease through haploinsufficiency, resulting in reduced levels of functional SynGAP protein. This mechanism is well established for many truncating and splice-disrupting variants. However, SynGAP is a multidomain protein containing several functionally distinct regions, including an N-terminal pleckstrin homology (PH) domain, a C2 domain, a Ras GTPase-activating (RasGAP) domain, and C-terminal regions involved in protein regulation and synaptic interactions. Variants affecting these regions may differ in their functional consequences, particularly when comparing variants expected to reduce protein expression with missense variants that alter specific amino acids within the protein.

Whether these molecular differences are reflected in distinct clinical outcomes remains insufficiently understood. Most clinical descriptions of *SYNGAP1*-DEE emphasize the overall disease phenotype, while relatively little is known about the relationship between individual protein domains and specific developmental abilities. Expressive language is particularly relevant in this context because it varies considerably between patients and represents a meaningful measure of functional development. Moreover, expressive language and functional communication are related but distinct abilities: individuals with limited speech may still communicate effectively through gestures, facial expressions, or other nonverbal means. Examining both dimensions may therefore provide a more complete picture of the clinical consequences of different *SYNGAP1* variants.

This distinction is becoming increasingly relevant as targeted therapies, including antisense oligonucleotides (ASOs) intended to increase functional SYNGAP1 expression, advance toward clinical development [5,12]. Such approaches aim to address the underlying molecular disorder rather than individual symptoms. However, it remains uncertain whether patients with different variant classes and affected protein domains have comparable baseline developmental profiles or will respond similarly to treatments that modify SYNGAP1 expression. A better understanding of genotype-specific language and communication outcomes could help inform patient stratification, treatment-arm balance, endpoint selection, and the interpretation of future clinical trials.

In the present study, we investigated the relationship between *SYNGAP1* variant location, variant class, and language outcomes using genetically verified, caregiver-reported data from the international PATRE SYNGAP1 registry. Rather than grouping patients according to overall clinical severity, we focused directly on current expressive language ability, acquisition of two-word phrases, and functional communication. Variants were assigned to predefined functional regions of the SynGAP protein, and associations were analyzed separately for missense and truncating/splicing variants. Our objective was to determine whether specific protein domains are associated with distinct language and communication profiles and whether these associations differ according to variant class, providing a clinically relevant foundation for genotype-informed research and future precision therapies.

## 2. Methods

### 2.1. Study Design and Registry Framework

This retrospective cross-sectional study used data from the international PATRE (PATient- Based phenotyping and evaluation of therapy for Rare Epilepsies) SYNGAP1 registry.

PATRE is a patient-centered digital platform developed to support structured phenotyping, treatment evaluation, and natural history research in rare developmental and epileptic encephalopathies [6].

The SYNGAP1 registry collects caregiver-reported information on molecular diagnosis, developmental milestones, epilepsy, behavior, language, communication, and everyday functioning. Data are collected through a multilingual digital application [8], allowing participation from families across different countries. The registry was initiated by the German SYNGAP1 patient organization in collaboration with its medical advisory board and is supported by a European network of patient organizations. The present study was conducted within the EURAS consortium (European network for neurodevelopmental RASopathies; Horizon Europe Grant No. 101080580) [7].

The analysis focused specifically on expressive language and functional communication in relation to SYNGAP1 variant class and protein domain. No composite disease-severity score or phenotype clustering was used.

### 2.2. Study Population and Inclusion Criteria

The registry dataset contained 440 individual participant records. For the present analysis, participants were eligible if their reported SYNGAP1 variant could be assigned to a protein position and classified as either a missense or truncating/splicing variant.

A total of 334 participants met these criteria, including 79 individuals with missense variants and 255 with truncating/splicing variants. Participants without sufficient variant information for protein-domain assignment or with variant types outside these two categories were excluded from the domain-based comparisons.

Availability of language and communication data varied between participants. Rather than requiring completion of all questionnaires, analyses were performed using the available observations for each outcome. Consequently, sample sizes differed across individual analyses. No missing clinical values were statistically imputed.

### 2.3. Variant Classification and Protein-Domain Assignment

SYNGAP1 variants were classified into two groups according to their reported molecular consequence. Truncating/splicing variants included nonsense, frameshift, canonical splice- site, and exonic variants with a possible splice effect. Missense variants were analyzed separately.

Variant positions were determined primarily from the reported protein-level HGVS annotation. When a protein position was unavailable, an approximate amino acid position was derived from the coding-DNA coordinate. The source of each positional assignment was retained for quality control.

Variants were assigned to seven predefined regions of the SynGAP protein:

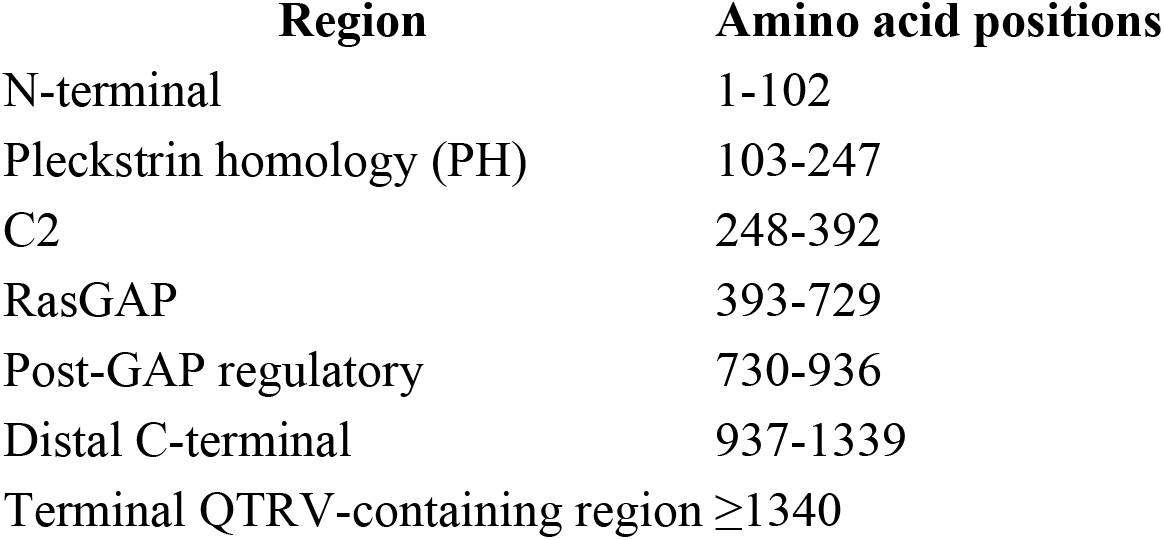

These regions were applied consistently across the entire protein. Each participant was assigned to one region according to the position of the reported variant.

For each region, language and communication outcomes were compared with those of participants carrying variants in all other protein regions. Comparisons were performed separately within the missense and truncating/splicing groups. This approach allowed regional associations to be evaluated without combining variant classes with potentially different molecular consequences.

### 2.4. Assessment of Expressive Language

The primary outcome was current expressive language ability, assessed using caregiver responses to structured questions within the PATRE registry.

The questionnaire first asked whether the patient’s current language abilities were age- appropriate. For patients whose language was not considered age-appropriate, an additional question assessed the highest level of expressive language ability.

Based on this branching structure, a five-level ordinal expressive language score was derived:

| **Level** | **Expressive language ability** |
|---|---|
| **1** | No understandable, meaningfully used words |
| **2** | Few single words (up to 50 words) |
| **3** | Numerous single words and very simple two-word phrases |
| **4** | Simple language with multiple words and complete sentences, but not age-appropriate |
| **5** | Age-appropriate language |

Higher scores indicated better expressive language ability.

Participants whose language was explicitly reported as age-appropriate were assigned level 5. For all other participants, the reported language category was retained. This approach accounted for the questionnaire’s branching logic without treating structurally unavailable responses as missing values.

The resulting score was used as an ordinal measure of current expressive language ability. It represents a registry-derived clinical classification rather than a separately validated standardized language assessment.

### 2.5. Developmental Language Milestones

As a secondary outcome, caregivers reported whether the patient had acquired the ability to speak two-word phrases.

This variable was analyzed as a binary developmental outcome (achieved versus not achieved). For participants who had acquired two-word phrases, additional questions documented whether this milestone had been reached by 24 months of age or later, as well as the reported age at acquisition in months.

The proportion of participants who had achieved two-word phrases was compared across protein regions. Age at acquisition and delayed acquisition after 24 months were evaluated only among participants who had achieved the milestone.

Participants who had not acquired two-word phrases were retained in the binary attainment analysis but were not assigned an artificial acquisition age.

### 2.6. Assessment of Functional Communication

Functional communication was assessed separately from expressive language using caregiver responses to the Communication Function Classification System (CFCS) decision questions.

The questionnaire evaluated whether the patient could effectively communicate as both sender and receiver, whether communication was effective with familiar and unfamiliar partners, and whether communication occurred at a comfortable pace.

Responses were used to derive CFCS levels I-V, ranging from effective communication with familiar and unfamiliar partners (level I) to severely limited communication effectiveness (level V).

For statistical analysis, CFCS levels were transformed into a functional communication score:

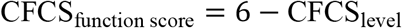

Higher scores therefore indicated better functional communication.

Additional caregiver-reported variables assessed the presence of communication difficulties and their impact on everyday functioning.

Expressive language and functional communication were analyzed as separate outcomes because effective communication may occur through speech, gestures, facial expressions, or other nonverbal means.

### 2.7. Statistical Analysis

All analyses were performed in R [10].

The primary analysis examined associations between protein-domain location and the five- level expressive language score. Each domain was compared with all remaining protein regions within the same variant class.

Descriptive results were summarized using medians and interquartile ranges for ordinal variables and proportions for binary outcomes.

Unadjusted comparisons were performed using Wilcoxon rank-sum tests for ordinal or continuous outcomes and Fisher’s exact tests for binary outcomes.

To account for potential differences in age between domain groups, the primary expressive language analysis additionally used proportional-odds ordinal logistic regression. Models included protein-domain membership and age as explanatory variables:

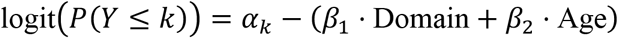

Age was treated as a continuous variable in years, calculated from the reported birth year and the analysis year (2026).

Odds ratios (ORs) and 95% confidence intervals were calculated for the domain effect. An OR greater than 1 indicated higher odds of belonging to a better expressive language category among participants with variants in the respective domain, while an OR below 1 indicated lower odds.

Secondary binary outcomes, including acquisition of two-word phrases, were assessed using Fisher’s exact tests and age-adjusted logistic regression where model estimation was feasible. Ordinal functional communication outcomes were analyzed using Wilcoxon tests and age- adjusted proportional-odds regression.

Models with insufficient complete observations, inadequate outcome variation, or unstable parameter estimates were not interpreted as providing reliable adjusted effects.

To account for multiple comparisons, Benjamini-Hochberg false discovery rate (FDR) correction was applied across the tested protein domains, separately for each variant class, outcome, and statistical test. An FDR-adjusted *q* < 0.05 was considered statistically significant. Results with 0.05 ≤ *q* < 0.10 were considered exploratory.

All statistical analyses were based on available observations without imputation of missing outcomes.

### 2.8. Ethics, Consent, and Data Availability

The study was conducted within the established PATRE research framework. Ethical approval was granted by the Ethics Committee of the Bavarian Medical Association (approval No. 21058; 25 August 2021).

Caregivers provided informed consent for registry participation and the use of de-identified data for research purposes. The present analysis used de-identified registry data.

## Data Availability

De-identified data underlying the findings of this study are not publicly available due to governance and privacy constraints in a rare disease registry context. Access to de-identified data may be considered upon reasonable request to the corresponding author and/or registry governance body, subject to scientific review, data use agreements, and applicable regulatory and ethical requirements.

## 3. Results

### 3.1. Study Cohort and Distribution of Variants Across SynGAP Domains

The registry dataset comprised 440 participants. Of these, 334 had sufficient molecular information to assign their SYNGAP1 variant to a protein position and one of the two predefined variant classes. The resulting study cohort included 255 participants with truncating/splicing variants and 79 with missense variants.

Variants were distributed across the predefined regions of the SynGAP protein, with substantial differences in the number of individuals represented within each domain and variant class. For the primary analysis of current expressive language, complete outcome and age data were available for 253 participants with truncating/splicing variants and 78 with missense variants.

Current expressive language varied considerably across the cohort, ranging from the absence of meaningful spoken words to age-appropriate language. Domain-specific differences were observed in both variant classes, with the strongest associations involving missense variants in the C2 domain and truncating/splicing variants in the PH domain (Figure 1).

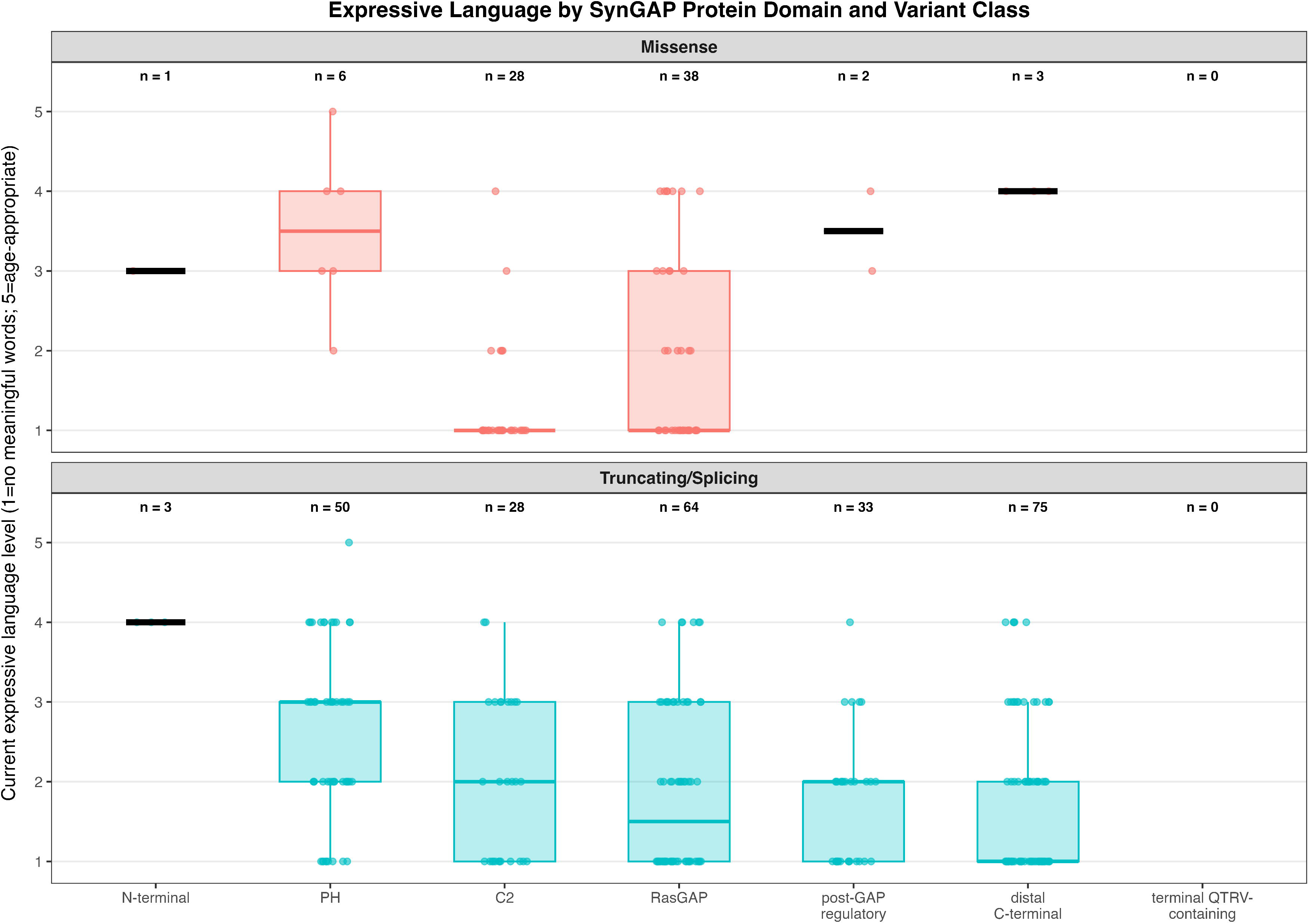

### 3.2. C2-Domain Missense Variants Are Associated With Poorer Expressive Language

Among participants with missense variants, those with variants in the C2 domain (aa248-392; n=28) had substantially poorer expressive language than individuals with missense variants elsewhere in the protein.

The median current expressive language score was 1 in the C2 group, corresponding to no meaningful spoken words, compared with 2 among participants with missense variants outside C2, corresponding to the use of a limited number of single words.

This difference was significant in the unadjusted Wilcoxon analysis (*p*=0.000379; FDR- adjusted *q*=0.0015).

The association remained significant after adjustment for age. In proportional-odds ordinal logistic regression, C2 missense variants were associated with markedly lower odds of belonging to a higher expressive language category (OR 0.16, 95% CI 0.06-0.46; *p*=0.000635; *q*=0.0025).

Thus, among individuals with missense variants, C2 localization was associated with substantially poorer current expressive language, independent of the age adjustment included in the model.

### 3.3. Reduced Acquisition of Two-Word Phrases and Functional Communication in C2 Missense Variants

The association between C2 missense variants and expressive language was also reflected in the developmental milestone of acquiring two-word phrases.

Only 2 of 28 participants (7.1%) with C2 missense variants had acquired two-word phrases, compared with 26 of 50 participants (52.0%) with missense variants outside the C2 domain.

This difference remained significant after adjustment for age (OR 0.07; *q*=0.0023).

Functional communication showed a similar pattern. Participants with C2 missense variants had lower CFCS function scores than those with other missense variants (median 2 vs. 3; Wilcoxon *p*=0.0025; *q*=0.0101).

The difference also remained significant after age adjustment (OR 0.23, 95% CI 0.09-0.56; *q*=0.0052).

Together, these findings show that the poorer expressive language associated with C2 missense variants was accompanied by lower attainment of an important language milestone and greater impairment in everyday functional communication.

### 3.4. PH-Domain Truncating/Splicing Variants Are Associated With Better Language Outcomes

In contrast to the unfavorable association observed for C2 missense variants, truncating/splicing variants in the PH domain (aa103-247; n=50) were associated with better expressive language.

The median current expressive language score was 3 among participants with PH-domain truncating/splicing variants, compared with 2 among individuals carrying truncating/splicing variants elsewhere in the protein.

This difference was significant in the unadjusted analysis (*p*=1.24×10⁻⁵; *q*=7.42×10⁻⁵).

Following adjustment for age, PH-domain truncating/splicing variants remained associated with higher odds of better expressive language (OR 3.52, 95% CI 1.99-6.24; *p*=1.59×10⁻⁵; *q*=9.54×10⁻⁵).

The same direction of association was observed for acquisition of two-word phrases. Among participants with truncating/splicing variants in the PH domain, 62.0% had acquired two-word phrases, compared with 31.5% of those with truncating/splicing variants outside PH.

This difference remained significant following age adjustment (OR 3.69; *q*<0.001).

These findings identify the PH domain as a region in which truncating/splicing variants are associated with relatively preserved expressive language compared with truncating/splicing variants in other parts of SynGAP.

### 3.5. Variant-Class Specificity and Findings in Other Protein Regions

The domain-associated language differences were not uniform across variant classes.

Importantly, the unfavorable language association observed for C2 missense variants was not observed for truncating/splicing variants within the same domain.

Among participants with C2 truncating/splicing variants (n=28), the median expressive language score was 2, identical to that of participants with truncating/splicing variants outside C2. Neither the unadjusted comparison (*p*=0.971) nor the age-adjusted analysis (OR 0.93, 95% CI 0.45-1.91; *p*=0.845) showed evidence of a corresponding association.

Similarly, no significant regional language association was identified for variants in the RasGAP domain in either variant class. The age-adjusted odds ratios were 0.94 for missense variants (*p*=0.889) and 0.73 for truncating/splicing variants (*p*=0.243).

Among truncating/splicing variants in the distal C-terminal region (aa937-1339; n=75), current expressive language was somewhat poorer than in the remaining truncating/splicing cohort (median 1 vs. 2). This association reached the predefined FDR threshold in the unadjusted analysis (*p*=0.0235; *q*=0.0470), but was weaker after age adjustment (OR 0.60, 95% CI 0.36-1.00; *p*=0.0498; *q*=0.0995).

Additional associations were observed in some sparsely represented domains, including N- terminal truncating/splicing variants and PH or distal C-terminal missense variants. However, these subgroups contained only three to six individuals, resulting in substantial uncertainty around their effect estimates. These findings were therefore considered exploratory.

Overall, the strongest and most consistently supported regional associations involved poorer expressive language and functional communication among C2 missense variants and better expressive language among PH-domain truncating/splicing variants.

## 4. Discussion

### 4.1. Principal Findings

This study identifies distinct associations between SynGAP protein domains, variant class, and language outcomes in a large international cohort of individuals with SYNGAP1-related developmental and epileptic encephalopathy. By focusing directly on expressive language and functional communication, rather than composite measures of overall disease severity, we found two particularly relevant patterns: missense variants within the C2 domain were associated with markedly poorer language and communication, whereas truncating/splicing variants within the PH domain were associated with better expressive language compared with variants of the same class located elsewhere in the protein.

The C2 association was particularly consistent. Among individuals with missense variants, C2 localization was associated with substantially lower odds of reaching a higher expressive language category after adjustment for age (OR 0.16, 95% CI 0.06-0.46). This difference was also reflected in developmental language milestones: only 7.1% of individuals with C2 missense variants had acquired two-word phrases, compared with 52.0% of those with missense variants outside C2. Functional communication, assessed using the CFCS, showed a similar association.

In contrast, truncating/splicing variants within the PH domain were associated with better expressive language (age-adjusted OR 3.52, 95% CI 1.99-6.24) and more frequent acquisition of two-word phrases. No comparable language association was observed for truncating/splicing variants within C2, while RasGAP variants showed no significant regional association in either variant class.

These findings suggest that protein location provides clinically relevant information beyond the broad distinction between missense and truncating/splicing variants. More importantly, they raise the possibility that variants affecting different functional regions of SynGAP may contribute to language impairment through different molecular mechanisms.

### 4.2. C2-Domain Missense Variants: Convergence of Clinical and Structural Evidence

The strongest adverse language association in our study involved missense variants within the C2 domain. This finding is particularly interesting in light of recent structural investigations into the molecular consequences of SynGAP missense variants.

In a study published in September 2026, Ali and colleagues investigated the interaction of SynGAP with Ras and Rap1b using molecular dynamics simulations, computational mutagenesis, and binding free-energy calculations. Their work provides a detailed structural framework for understanding how individual missense substitutions may interfere with SynGAP function [13].

The authors propose that SynGAP regulates Ras through a relatively conventional RasGAP mechanism, whereas its interaction with Rap1b involves a distinct, Plexin-like mechanism. In the latter model, the C2 domain contributes to stabilizing the conformation of Rap-Switch II required for efficient GTP hydrolysis. This implies that the C2 domain is not merely an accessory structural element but may participate directly in the organization of the SynGAP- GTPase interaction.

Importantly, the authors identified several substitutions affecting the C2 region that were predicted to destabilize interactions with Ras or Rap. These included R272W and E360K, as well as substitutions involving Arg401 at the structurally defined C2 interaction interface. The predicted effects included disruption of electrostatic contacts, reduced stability of the protein- protein interface, and changes in the positioning of residues involved in catalytic regulation.

These structural findings provide a plausible molecular context for the pronounced language impairment observed in our C2 missense subgroup. While our registry data cannot establish the precise functional consequences of individual variants, the association between C2 localization and impaired expressive language is consistent with the possibility that alterations in this domain interfere with functions extending beyond a simple reduction in total SynGAP abundance.

The potential contribution of Rap signaling is particularly intriguing. Ali et al. propose that the C2 domain plays a critical role in stabilizing the open Rap-Switch II conformation required for efficient SynGAP-mediated Rap inactivation. Their models further suggest that certain substitutions may affect Ras and Rap interactions differently, raising the possibility that missense variants can alter specific aspects of SynGAP signaling rather than producing an equivalent reduction in all protein functions.

Such differences could contribute to clinical heterogeneity, although the relationship between altered Ras/Rap regulation and expressive language development remains to be established experimentally.

The relevance of the C2 domain may also extend beyond catalytic activity. Its proposed contribution to membrane interactions and the structural organization of SynGAP-GTPase complexes suggests that missense variants could interfere with protein localization, molecular interactions, or the organization of postsynaptic signaling complexes.

This distinction is important because recent experimental studies have demonstrated that the catalytic and structural functions of SynGAP are not necessarily equivalent in their contribution to neurodevelopmental phenotypes [15]. Consequently, the adverse language association observed in our C2 subgroup should not be attributed exclusively to impaired Ras/Rap GTPase activity.

Our clinical findings and the structural models of Ali et al. therefore provide complementary evidence: the former identifies a domain-associated clinical phenotype, while the latter suggests specific molecular mechanisms through which C2-region substitutions might alter SynGAP function.

Further studies combining patient-derived cellular models, direct measurements of protein expression and stability, and functional analysis of Ras/Rap signaling will be required to determine which of these mechanisms contribute to the observed language phenotype.

### 4.3. PH-Domain Truncating/Splicing Variants and Relative Preservation of Language

A second important finding was the association between truncating/splicing variants in the PH domain and relatively better expressive language.

Participants with PH-domain truncating/splicing variants were more likely to demonstrate higher current language abilities and had more frequently acquired two-word phrases than individuals with truncating/splicing variants elsewhere in the protein.

This observation suggests that the clinical consequences of variants classified as truncating or splice-disrupting may not be uniform across the SynGAP protein.

Haploinsufficiency remains the principal established disease mechanism for many pathogenic SYNGAP1 variants. Premature termination codons and certain splice-disrupting variants are expected to reduce the abundance of functional SynGAP, often through nonsense-mediated mRNA decay (NMD). However, the consequences of individual variants may depend on their transcript context, position relative to exon junctions, alternative splicing, and the expression of different SynGAP isoforms.

These factors could potentially contribute to differences in functional protein availability or the production of residual protein products.

One possible explanation for the favorable association observed in the PH-domain subgroup is that variants in this region have different transcript-level consequences from variants in other parts of the gene. Alternatively, the observed effect could reflect differences in isoform expression or other molecular features not captured by broad variant-consequence categories.

However, our findings do not establish that PH-domain truncating variants undergo more efficient NMD or preserve specific SynGAP functions. NMD is not determined by protein- domain membership alone, and many truncating variants outside PH are also expected to undergo transcript degradation.

Furthermore, the favorable language association should not be interpreted as evidence that PH-domain truncating variants produce a generally mild form of SYNGAP1-DEE. Our analysis addresses expressive language and communication, not overall disease severity or longitudinal developmental trajectories.

The biological basis of this association therefore remains uncertain. Transcript-specific annotation, RNA expression analyses, and studies of protein abundance and isoform composition may help determine whether PH-domain truncating/splicing variants represent a functionally distinct subgroup.

### 4.4. Implications for the Haploinsufficiency Model

Our results support a more differentiated understanding of genotype-phenotype relationships in SYNGAP1-DEE.

Haploinsufficiency is well established as a major disease mechanism, particularly for pathogenic variants that reduce functional SynGAP expression. However, this mechanism does not necessarily imply that all pathogenic variants produce identical molecular consequences or equivalent clinical outcomes.

The present findings suggest that both variant class and protein location are relevant to expressive language development.

The unfavorable association observed for C2 missense variants, together with the absence of a corresponding association among C2 truncating/splicing variants, is particularly noteworthy. This difference is compatible with the possibility that at least some missense substitutions alter specific protein functions in ways that are not reproduced by truncating variants.

Such effects might include impaired catalytic regulation, altered interactions with signaling partners, changes in protein stability or localization, or interference with postsynaptic protein complexes.

These possibilities are consistent with the structural diversity of missense effects proposed by Ali et al. [13], who demonstrated through molecular modelling that different substitutions may affect SynGAP-GTPase interactions through distinct mechanisms.

Importantly, neither the present clinical data nor the available structural models establish dominant-negative activity or toxic gain-of-function as a general mechanism for C2 missense variants. Severe phenotypes may also result from substantial loss of specific protein functions without requiring an active interference mechanism.

Moreover, although the domain associations differed descriptively between variant classes, a formal domain-by-variant-class interaction analysis would be required to establish whether these effects differ statistically between missense and truncating/splicing variants.

Our findings therefore do not challenge haploinsufficiency as a central mechanism of SYNGAP1-related disease. Rather, they indicate that a uniform haploinsufficiency model may be insufficient to account for the observed variation in language and communication outcomes across different molecular variant groups.

### 4.5. Clinical Implications and Relevance for Upcoming ASO Trials

The identification of domain-associated differences in language and communication may have important implications for clinical assessment and the development of targeted therapies.

Expressive language is a particularly relevant clinical outcome in SYNGAP1-DEE because the ability to communicate meaningfully affects everyday functioning, social participation, and caregiver-patient interactions. The substantial differences observed between variant- defined subgroups suggest that molecular genotype may contribute to the variability of these abilities.

For clinical counseling, information about variant class and protein location could potentially complement existing assessments when discussing expected developmental abilities with families. However, the substantial variability among individuals and the cross-sectional nature of the present study preclude deterministic predictions for individual patients.

The translational implications are especially relevant as antisense oligonucleotides and other approaches intended to increase functional SYNGAP1 expression move toward clinical evaluation.

Such therapies are generally designed around the concept that increasing functional SynGAP availability may improve disease-related outcomes. However, the molecular consequences of different pathogenic variants may influence both baseline clinical characteristics and the biological effects of a given therapeutic strategy.

For variants associated with transcript degradation and reduced functional protein expression, increasing expression from the remaining functional allele may be a biologically appropriate approach.

In contrast, some missense variants may produce altered proteins with residual, selectively impaired, or potentially interfering functions. Depending on the mechanism of the therapeutic approach, increasing gene expression might affect wild-type and mutant alleles differently.

The consequences of such treatment cannot be inferred from the present study and require variant-specific functional investigation.

Our findings therefore support considering variant class and protein domain when characterizing participants in future ASO trials. Such information could be relevant for cohort composition, treatment-arm balance, subgroup analyses, and interpretation of developmental outcomes.

The marked baseline differences in expressive language are particularly important for endpoint selection. A participant without meaningful spoken words and a participant already using sentences may have substantially different opportunities to demonstrate improvement on a categorical language scale. Baseline language ability, age, and communication function should therefore be considered when evaluating the sensitivity and clinical relevance of language-related endpoints.

Functional communication may provide complementary information, particularly for individuals whose expressive speech remains severely limited but who communicate through gestures, facial expressions, or alternative communication methods.

Ultimately, genotype-informed stratification may help distinguish differences in baseline disease expression from potential treatment-related changes. However, the present findings do not establish that any subgroup will respond more favorably or less favorably to ASO treatment. Prospective clinical studies and functional investigations are needed before protein- domain location can be considered a predictive biomarker of therapeutic response.

### 4.6. Strengths and Limitations

A major strength of this study is the use of a large international registry cohort with structured caregiver-reported language and communication data linked to molecular variant information.

Unlike analyses based on multidimensional severity composites, the present study focused on directly interpretable clinical outcomes. Expressive language ability, acquisition of two-word phrases, and functional communication were evaluated separately, allowing the consistency of regional associations to be assessed across related but distinct measures.

The systematic analysis of predefined protein regions, performed separately for missense and truncating/splicing variants, also reduced the dependence on arbitrarily selected protein intervals. Age-adjusted models and false discovery rate correction were used to address potential age differences and multiple comparisons.

#### Several limitations warrant consideration

First, the study was retrospective and cross-sectional. Current language ability and retrospectively reported developmental milestones cannot establish individual developmental trajectories or predict future language acquisition.

Second, the language and communication measures were caregiver-reported. Although they capture clinically meaningful everyday abilities, they are not equivalent to standardized clinician-administered developmental assessments. The five-level expressive language score was derived from the registry questionnaire and has not been independently validated as a standardized outcome measure. Similarly, CFCS levels were reconstructed from caregiver responses rather than established through a dedicated clinician-administered classification process.

Third, the age-adjusted analyses used age estimated from birth year and the analysis year. Differences between this estimate and the actual age at questionnaire completion may introduce residual measurement error. In addition, the assumption of a linear age effect and the proportional-odds assumption require further assessment, particularly because language acquisition is strongly age-dependent during early childhood.

Fourth, the distribution of variants was uneven across protein regions. Several domains contained only small numbers of individuals, especially within the missense group. Recurrent variants may also contribute disproportionately to observed regional associations, and the extent to which individual substitutions drive the C2 association requires further investigation.

Fifth, domain assignment was based on a predefined protein annotation. Boundaries between functional domains may vary between annotations or structural definitions, and SynGAP isoforms can differ in their sequence organization. These considerations are relevant when comparing our clinically assigned protein positions with structural models derived from specific isoforms.

Sixth, the study does not directly assess the molecular consequences of individual variants. We did not measure transcript abundance, NMD efficiency, protein stability, subcellular localization, or Ras/Rap signaling. Although the work of Ali et al. [13] provides a plausible structural framework, its computational predictions do not establish the functional consequences of the variants represented in our registry cohort.

Finally, additional genetic, environmental, therapeutic, and ascertainment-related factors may contribute to language outcomes. The observational design does not permit causal attribution of language impairment to particular molecular pathways or protein domains.

### 4.7. Future Directions

The present findings identify several opportunities for further investigation.

A particularly relevant next step would be to combine detailed clinical phenotyping with functional characterization of recurrent and representative C2 missense variants. Patient- derived neuronal models could be used to examine protein abundance, synaptic localization, interactions with Ras and Rap, and downstream signaling.

The structural predictions of Ali et al. [13] provide a framework for selecting molecular interactions and functional mechanisms for such investigations. Comparing experimentally measured effects with language and communication outcomes may help determine whether particular molecular abnormalities are associated with more severe neurodevelopmental phenotypes.

Complementary evidence was recently provided by Miotto et al. [14], who investigated the membrane-binding dynamics of the SynGAP C2 domain using molecular dynamics simulations. Their findings suggest that C2 missense variants can alter membrane interactions and lateral diffusion without substantially disrupting the overall protein fold. These observations provide an additional mechanistic perspective on our clinical findings. Together with the structural models of Ali et al. [13], they suggest that C2 missense variants may interfere with multiple aspects of SynGAP function, including membrane-associated dynamics and Ras/Rap-GTPase interactions. Whether these molecular alterations contribute to the pronounced language impairment observed in our cohort remains to be established experimentally.

Similarly, transcript-specific analyses of PH-domain truncating/splicing variants could clarify whether differences in RNA processing, isoform expression, or residual protein production contribute to the relatively favorable language association observed in this subgroup.

Longitudinal follow-up will also be essential. Repeated language and communication assessments within the PATRE registry could establish whether the domain-associated differences observed cross-sectionally correspond to distinct developmental trajectories, rates of skill acquisition, or changes in functional communication over time.

These investigations could ultimately support the development of biologically informed patient subgroups and more sensitive clinical outcome measures for future targeted-treatment studies.

Taken together, our findings provide clinical evidence that expressive language and functional communication in SYNGAP1-DEE are associated with variant class and protein-domain location. The convergence between the adverse C2 missense phenotype and recent structural models of altered SynGAP-GTPase interactions offers a compelling basis for further mechanistic research. As targeted therapies advance, integrating clinical phenotyping with molecular characterization may become increasingly important for understanding disease heterogeneity and designing genotype-informed clinical trials.

## 5. Conclusion

Our findings demonstrate that expressive language and functional communication in SYNGAP1-DEE are associated with both protein domain and variant class. C2-domain missense variants were linked to markedly poorer language outcomes, whereas truncating/splicing variants in the PH domain were associated with relatively preserved expressive language. Together with emerging structural evidence, these findings suggest that SYNGAP1-related disease cannot be fully understood through a uniform haploinsufficiency model alone. As targeted therapies, including antisense oligonucleotides, advance toward clinical trials, considering variant class and protein location may be important for patient stratification, endpoint selection, and interpretation of treatment outcomes.

## Data Availability

De-identified data underlying the findings of this study are not publicly available due to
governance and privacy constraints in a rare disease registry context. Access to de-identified
data may be considered upon reasonable request to the corresponding author and/or registry
governance body, subject to scientific review, data use agreements, and applicable regulatory
and ethical requirements.

